# Using Synthesised Narrative Exploration to generate contextually grounded insights for HIV prevention, testing and care programming in Mozambique

**DOI:** 10.64898/2026.03.23.26349138

**Authors:** Pam Baatsen, Tavares Madede, Nwanneka Okere, Noor Tromp, Mari Luntamo

## Abstract

Evidence-informed situation analyses are critical for HIV programming, yet primary data collection is often resource-intensive and time-consuming. Approaches that leverage existing evidence while generating locally grounded insights are needed to inform context-appropriate interventions.

We applied the Synthesised Narrative Exploration (SNE) approach in two districts in Inhambane province, southern Mozambique, to inform an HIV project through assessing key dynamics affecting HIV prevention, treatment, and care for women and children. SNE combines structured synthesis of existing country-specific literature with qualitative validation and contextualisation through focus group discussions and semi-structured interviews. Findings from 16 peer-reviewed studies and two national reports were synthesised into short narratives and discussed with 83 community members and key stakeholders. Qualitative data were thematically analysed and validated through community and stakeholder consultations.

Study participants confirmed many well-documented barriers across the maternal and infant HIV care cascade and added local insights. Known pre-pregnancy barriers such as limited preconception HIV testing and low ART uptake were confirmed, while strong fertility expectations, low levels of pregnancy planning, gendered decision-making, and fear of partner abandonment following HIV status disclosure were highlighted as factors shaping engagement with HIV services. The participants confirmed barriers during pregnancy reported in earlier studies, including delayed antenatal care initiation and limited male partner involvement, and added the role of household power hierarchies, particularly the influence of mothers-in-law, in shaping HIV testing, disclosure, and ART adherence. Widely documented postpartum disengagement from care was explained by difficulties sustaining non-disclosure and the loss of socially acceptable reasons for continued clinic attendance. The participants proposed locally grounded strategies to address the barriers, including strengthened couple counselling and engagement of influential family members such as mothers-in-law.

By integrating existing evidence in a qualitative enquiry, SNE enables efficient generation of contextually rich insights directly relevant to intervention design, offering an approach for strengthening HIV programmes.

## Introduction

Mozambique is one of the countries most affected by the HIV epidemic. In 2021, the year in which data for this study were collected, adult HIV prevalence was estimated at 12.4%, with a substantially higher prevalence among women (15.1%) than men (9.3%). In the same year, an estimated 2.4 million people of all ages were living with HIV, including approximately 190,000 children between 0–14 years. Nearly one in five people living with HIV (19%) were not aware of their HIV status. In addition, one third (33%) of all people estimated to be living with HIV were not receiving antiretroviral therapy (ART). The treatment gap was particularly pronounced among children living with HIV, 54% of whom were not on ART in 2021 [1].

Despite progress in HIV service delivery, substantial gaps remained in prevention of mother-to-child transmission (PMTCT) and paediatric HIV care. Prior research in Mozambique documented consistently low ART uptake before pregnancy [2–4], delayed initiation of antenatal care [5–7], and suboptimal retention in HIV care during and after pregnancy [8,9]. Limited uptake of early infant diagnosis, and poor ART adherence among children were also reported [10]. Together, these gaps reflected persistent vulnerabilities across the maternal and child HIV care cascade. However, while the above-mentioned service level and behavioural barriers were well documented, less attention had been given to how local social norms, family dynamics, and health system realities shaped care-seeking behaviours, HIV status disclosure practices and treatment adherence across the continuum of maternal and child health, from pre-pregnancy, through pregnancy, delivery, infancy and childhood.

For this reason, there was a need for an approach that could build on existing evidence while generating context-specific insights to inform locally appropriate programmatic responses, without relying solely on resource-intensive and time-consuming primary data collection. Rapid reviews and rapid assessment approaches have been developed to support timely evidence-informed decision-making [11,12]. These approaches take multiple forms, including rapid epidemiological assessments, rapid appraisal methods, and rapid assessment procedures [13,14], and aim to balance speed, adaptability, and analytical depth in response to urgent public health needs [15]. However, while these approaches are valuable for rapidly summarising situations, their emphasis on timeliness and pragmatism can limit the extent to which they systematically engage with the full body of existing empirical evidence or how deeply they explore the context-specific social dynamics, lived experiences, and explanatory mechanisms that shape health behaviours and patterns of service utilisation.

The Synthesised Narrative Exploration (SNE) approach was developed to address this gap. Building on the principles of rapid assessments, SNE integrates existing country-level evidence directly into qualitative data collection. Findings from prior research are synthesised into short narratives that are presented to community members and stakeholders for validation, refinement, and contextualisation. By embedding synthesised evidence directly into study participant discussions, the approach seeks to bridge the gap between established research and local realities relevant for programme design.

In this study, we applied the SNE approach in Jangamo and Massinga districts in southern Mozambique to examine the social, behavioural, and structural determinants shaping HIV prevention, treatment, and care among women and children in the implementation areas of an HIV project. The objective was to generate context-specific evidence to strengthen HIV programming by grounding intervention design in both existing research and community perspectives.

## Materials and methods

### Study setting

The study was conducted in Jangamo and Massinga districts in Inhambane province, southern Mozambique, within the catchment area of one primary health facility in each district, to inform the Kusingata HIV project implemented by N’weti, a Mozambican non-governmental organisation. The project aimed to improve uptake of and retention in HIV services through a combination of community- and facility-based strategies, with a focus on children and mothers living with or affected by HIV in rural communities with limited access to health services and persistent gaps in maternal and child HIV care.

### Study design

This study employed the SNE qualitative research approach (S1 Appendix), developed by the first author as a structured method for integrating existing empirical evidence into stakeholder-engaged qualitative enquiry. The study followed a predefined process, including selection of a conceptual framework, a structured literature review, synthesis of findings into short narratives with guiding prompts, qualitative data collection, thematic analysis, and stakeholder validation.

This study was guided by the adapted socioecological model (SEM), which conceptualises health behaviours as shaped by interacting influences at individual, interpersonal, institutional, community, and policy levels [16]. The SEM has been widely applied in HIV and sexual and reproductive health research to assess how individual knowledge and beliefs, interpersonal relationships, service delivery systems, and social contexts jointly influence health-related decision making and engagement with care [17, 18].

The framework was combined with key stages of the maternal and child health continuum (pre-pregnancy, pregnancy, and postpartum) [19, 20] to explore how multilevel factors influence HIV testing, disclosure, treatment initiation, retention in care, adherence to treatment, and early infant diagnosis. At the individual level, we reviewed knowledge, perceptions, and practices related to fertility and health-seeking behaviours related to PMTCT. Interpersonal influences encompassed partner and family dynamics, including HIV status disclosure, male partner involvement, and household decision-making. Community-level influences captured the role of social norms and community structures in supporting or constraining engagement with HIV services. Institutional factors reflected the role of health workers and HIV-related services in shaping access to and continuity of care.

A structured literature review (S2 Appendix) was conducted to identify existing evidence in Mozambique that was relevant to the conceptual framework and the study objective. Searches were performed in Google Scholar using combinations of geographic and thematic terms derived from the conceptual framework. Titles and abstracts were screened for relevance, followed by full-text review. Peer-reviewed articles published between January 2010 and March 2020 in English or Portuguese were eligible. Grey literature, including national policy and programme reports, was also reviewed. Studies were included if they addressed barriers or enablers related to the thematic domains of the conceptual framework. In total, 16 peer-reviewed articles and two national reports met the inclusion criteria.

Key findings were extracted in a study matrix aligned with the conceptual framework. For each thematic domain, evidence was synthesised into concise and accessible narrative summaries. These summaries were then accompanied by open-ended discussion questions designed to encourage participants to confirm, nuance, or challenge the evidence and explain how it applied within their local context. The narratives served as entry points for discussion rather than statements of fact. Additional literature, including more recent studies and research from related sub-Saharan African contexts, was consulted to contextualise the findings and support interpretation in the introduction and discussion sections of this manuscript. These sources were not used to construct the SNE narratives but to situate the study findings within the broader evidence base.

### Participant recruitment and data collection

Qualitative data were collected in July 2021 through 11 focus group discussions (FGDs) with 64 participants, and 19 individual semi-structured interviews (SSIs), yielding a total sample of 83 participants. Data collection was conducted by one of the authors, a Mozambican national, together with a female Mozambican colleague, both based in Maputo, and two locally recruited and trained research assistants from the study districts. Interviews and discussions were conducted in Portuguese or the relevant local language, according to the participants’ preference. Participants were purposively selected to capture diverse perspectives across age, sex, HIV status, caregiving roles, and professional or community positions. Potential participants were identified with the assistance of local stakeholders, including community leaders and health workers, while recruitment and invitations to participate were conducted directly by the study team. The sample included young (15-19 years) and adult (20 years and above) women living with HIV who had children, men living with HIV who had children, women and men irrespective of HIV status who had children, women without children, healthcare providers, community leaders, community actors and civil society representatives working with HIV or related services. The FGDs were stratified by age and sex to facilitate open discussion. The narratives developed from the literature review were presented during FGDs and SSIs, followed by facilitated probing to explore similarities, differences, and underlying explanations. Individuals residing outside the study area, those with prior exposure to N’weti’s interventions, and those with conditions impairing their ability to provide informed consent or meaningfully participate, were excluded. All sessions were audio-recorded with participant consent. The study tool is provided in S3 Appendix.

### Data management and analysis

Audio recordings were transcribed verbatim in Portuguese, with direct translation from local languages during the transcription process where required. Transcripts were coded in Portuguese by the second and first author using a thematic framework derived from the conceptual framework and the narrative discussion guides, with additional inductive codes added to capture emerging themes. The two coders independently reviewed an initial set of transcripts and compared coding to ensure consistency, resolving discrepancies through discussion to build consensus and refine the coding framework. Coding and data management were conducted in NVivo 12 software and coded data were organised by thematic domain within the programme. Thematically organised data were subsequently analysed to identify key barriers, enablers, and contextual dynamics influencing HIV service utilisation during the pre-pregnancy, pregnancy, and postpartum periods. Attention was paid to points of convergence with the existing literature, contextual divergences, and newly identified or previously underexplored mechanisms shaping care-seeking, disclosure, and engagement in care and treatment. Findings were then synthesised and written up in English.

### Validation of findings

To enhance credibility and interpretation, the preliminary findings were validated through structured feedback sessions with community members and other stakeholders. Community validation meetings were held in February 2022, followed by an online stakeholder consultation in April 2022. Stakeholder feedback informed prioritisation of findings and refinement of recommendations, including participant-generated suggestions, to enhance their relevance and feasibility for programme design.

### Ethical considerations

Ethical approval was obtained from the Bioethics Committee of Eduardo Mondlane University, Maputo, Mozambique in May 2021, and from the Mozambique Ministry of Health in July 2021 (S4 Appendix). All participants provided written informed consent prior to participation. For participants under 18 years of age, written informed assent was obtained, and written informed consent was provided by a trusted adult acting as a guardian.

The FGDs were held in private rooms within community venues or health facilities. Interviews were conducted in private settings. Audio recordings and transcripts were securely stored, transcripts were de-identified, and all information presented anonymised.

## Results

Because the SNE approach integrates prior research directly into qualitative data collection, the results provided here reflect both key themes synthesised from the literature, and how participants validated, contextualised, or challenged those themes. Appendix S5 provides an overview of synthesised evidence from prior studies and how participants validated, contextualised, and expanded these findings.

Overall, participants confirmed many patterns described in previous studies while offering additional explanations and locally specific dynamics shaping HIV prevention, testing, treatment, and care for women and children. Findings are presented across key areas of the maternal and child health continuum, before pregnancy, during pregnancy and during the postpartum period.

### Before pregnancy

#### Fertility expectations and planning for pregnancy

Synthesised evidence from Mozambique indicated that pregnancies were frequently unplanned and strongly shaped by social expectations of motherhood [2, 21]. Participants in this study affirmed these norms, describing motherhood as central to a woman’s social identity, relationship stability, community respect, while also linking it to economic security and family lineage. Participants also described adolescent pregnancy as common and highlighted conflicting views around contraception, particularly regarding young people’s use of contraceptive methods prior to having children. Participants emphasised that the importance of having children did not differ between people living with HIV and those who were not infected. Health workers reported that they were not aware of cases where individuals had decided not to have children because of their HIV status, suggesting that strong reproductive expectations persist regardless of HIV status.

In contrast to earlier findings suggesting limited engagement with preconception care among women living with HIV, several women interviewed in this study described being motivated to seek guidance from health workers before becoming pregnant. Some stakeholders nevertheless suggested that opportunities for preconception counselling could be strengthened. They noted that health providers could more systematically ask women receiving ART whether they intend to become pregnant, thereby creating additional opportunities for counselling and preparation prior to conception.

#### Barriers to HIV testing and ART uptake before pregnancy

Prior research suggested that HIV testing before pregnancy was uncommon, with many women learning their status and initiating ART only during pregnancy [2,21,7,22]. Participants in this study confirmed that pre-pregnancy testing was rare and explained that people generally seek care only when ill, reducing opportunities for early diagnosis and treatment.

Couple testing was described as particularly sensitive. Participants explained that couples rarely tested together because of fears that discordant results could create conflict or threaten relationship stability. Both men and women noted that if one partner tested HIV positive while the other tested negative, the positive partner might be accused of infidelity, potentially leading to blame, disputes within the household, or relationship breakdown.

> *“Many times, we as men, think that taking the [HIV] test together with our wife could result in getting a positive [HIV] result and our relationship then ends.”* (Man, FGD, Jangamo)

Fear of disclosure within relationships also shaped ART uptake before pregnancy. Participants described how women who had not disclosed their HIV status often hid medication, avoided or invented alternative reasons for clinic visits, or discarded pills to prevent suspicion.

> *“Other women…throw them [the ART pills] away on the way home because the husband cannot know that she is taking antiretrovirals.”* (Woman living with HIV, SSI, Jangamo)

These accounts illustrate how concerns about blame, relationship conflict, or accusations of infidelity influenced women’s ability to engage openly with HIV services and the constant negotiation required to be able to adhere to treatment secretly. As a result, some women living with HIV were more concerned with protecting their relationships than their own health, and therefore did not consistently adhere to treatment.

Participants also described practical challenges related to ART use. Some reported that when they first started treatment they had not been fully informed about potential side effects, which initially caused anxiety and reluctance to continue treatment. However, many noted that these concerns diminished over time as they became accustomed to the medication and followed advice to take the pills with food. Women living with HIV frequently emphasised the importance of strict adherence, explaining that consistent treatment allowed them to maintain good health and reduce the risk of mother-to-child transmission.

Food insecurity was also described as a barrier to consistent ART use. Participants explained that many households depend on small-scale farming for their livelihoods and often eat only one substantial meal late in the morning. Taking medication on an empty stomach was said to cause nausea or weakness, making adherence more difficult for some individuals. Seasonal food shortages were also mentioned as influencing treatment continuity, with participants explaining that during lean months some people skipped doses, delayed clinic visits, or abandoned treatment altogether.

Together, these findings suggested that engagement in HIV testing and treatment before pregnancy was constrained by gendered power relations, fear of disclosure, and livelihood constraints. Many women had their first sustained point of contact with HIV services, including HIV testing and access to treatment when needed, only when they became pregnant.

### During pregnancy

#### Entry into care: late antenatal attendance and repeat testing

The literature highlights delayed antenatal care (ANC) initiation as a barrier to early HIV testing and ART initiation during pregnancy [7,3]. The participants of this study confirmed that many women presented late for ANC, particularly when pregnancies were unintended. Reasons included hiding pregnancies from parents, fear of HIV-testing as part of the ANC protocol, concerns about not being treated well by healthcare staff, and long distance to the health facilities especially in remote areas.

> *“Most cases are those unwanted pregnancies, the person is afraid to reveal that they are pregnant. So, they keep guarding, hiding, wearing big blouses…”*. (Health provider, Jangamo, SSI)

Participants also suggested that unequal access to health information contributed to delayed engagement with ANC services. Male participants in a FGD in Jangamo district noted that women with higher levels of education were more likely to receive and understand health information and therefore more likely to seek care early. They further explained that health information campaigns were often perceived as being concentrated in towns and larger villages, leaving poorer and less educated women in more remote rural areas with limited exposure to these messages.

Repeat HIV testing among women who had previously received an HIV-positive test result but had subsequently discontinued treatment [2,21], was also reported by participants. They explained that treatment discontinuation was often linked to non-disclosure of their HIV status to their partner, as well as misconceptions about HIV and its treatment. According to these participants, some women believed that HIV could disappear with treatment, while others perceived HIV-related care as necessary only during pregnancy to protect the child, making long-term treatment appear unnecessary. When they got pregnant, these women were tested for HIV again as part of ANC.

#### Relational and family dynamics shaping HIV testing and ART use

Once women entered care, their continued engagement was shaped not only by service access but also by relationship and family dynamics. According to previous studies, male partner involvement during pregnancy is associated with improved HIV testing uptake and ART adherence, yet remained limited in many settings [22,23]. Participants in this study agreed that most women in the implementation areas of Jangamo and Massinga districts attended ANC alone, with many men working abroad. Other men avoided accompanying their partners due to fear of ridicule from peers, as men who attend the health facility with their wives may be perceived as being under their wives’ control, which conflicts with prevailing expectations of male authority and independence. Yet other men avoided attending ANC with their wives due to the protocol that includes an HIV test for both partners.

Participants emphasised that partner awareness and support could significantly facilitate treatment adherence. Women explained that when their partners were aware of their HIV status, it became much easier to take ART consistently, as they no longer needed to hide the medication or conceal clinic visits. In contrast, women who had not disclosed their status often faced difficulties taking medication openly within the household.

Participants also shared that women without partners and those in polygamous relationships faced additional challenges due to limited social and economic support, as well as more complex disclosure and decision-making dynamics around HIV testing and treatment.

One previous study indicated that mothers-in-law influence ART use, with some associating it with impurity or infertility, occasionally leading to reactions like expulsion of the daughter-in-law from the household [2]. The study participants confirmed this and reported cases where mothers-in-law discouraged ART use or blamed daughters-in-law for introducing HIV into the family.

> *“It happens a lot, the mother-in-law sends her daughter-in-law away because she knows she is taking antiretrovirals; and at some point she claims that her daughter-in-law is the one who brought the disease, all this without knowing that her son is also HIV positive.”* (Adolescent woman, FGD, Massinga)

Participants also mentioned misconceptions among some mothers-in-law about ART, including the belief that the pills could harm the unborn child or even cause the baby to die.

Participants in this study said that some women questioned the need for ART when they felt physically well, a pattern also noted in previous studies [9, 24]. The participants explained that this was sometimes linked to disbelief in the diagnosis, particularly among women who first learned their HIV status during pregnancy while still feeling healthy. Fear of stigma also affected adherence, with some, especially younger women, avoiding collecting medication from health facilities or mobile brigades because they feared being seen by neighbours or community members. Others expressed concerns about the perceived side effects of ART, including fears that taking medication daily could cause them to become visibly thin. Participants also noted that some women felt less motivated to adhere to treatment when the pregnancy was unintended.

Participants added that abandonment by the partner and social isolation in such cases further discouraged ART adherence, particularly among adolescents. At the same time, protecting the unborn child was described as a strong motivator for adherence. Counselling during ANC and one-stop service models was reported to reinforce this motivation by highlighting the role of treatment in preventing HIV transmission to the baby and encouraging some women to restart ART after interruption.

These findings illustrate how pregnancy created both an opportunity and a vulnerability: while concern for the unborn child motivated engagement in care, stigma, relationship instability, and family hierarchies undermined sustained treatment and HIV status disclosure. These dynamics did not end with delivery, rather, the postpartum period introduced new challenges that further affected continuity of care for both mothers and infants.

### Postpartum period

#### Disengagement from maternal HIV care after delivery

Loss to follow-up after delivery has been widely documented in PMTCT programmes [9,25]. The participants of this study confirmed that many women disengaged from care postpartum and provided additional insight into why this occurred. A key theme was that pregnancy had provided a socially acceptable reason to attend health services, which diminished after childbirth.

> *“ [After the baby is born] she no longer has a justification for her family or partner to continue going to the health facility every month.”* (Civil society stakeholder, SSI, Massinga)

When HIV status had not been disclosed to partners or other family members at home, continued clinic visits became difficult to explain, contributing to treatment interruption. Health providers also noted that some women living with HIV were more concerned about the health of their child than about their own health. According to these providers, some women viewed their primary responsibility as ensuring that the child remained HIV negative during pregnancy and breastfeeding, while considering themselves already ill and therefore placing less emphasis on their own continued care. As a result, some women adhered to ART during pregnancy and possibly during breastfeeding but discontinued treatment once the child was born or after weaning.

Health workers further explained that treatment interruption after weaning was relatively common. While some women disengaged because of the distance to health facilities or fear of stigma, others were described as stopping treatment once they felt the immediate risk of HIV transmission to the child had passed.

These patterns of maternal disengagement were closely linked to how caregivers understood and prioritised infant HIV testing and follow-up.

#### Infant testing, misconceptions, and family influence

Previous studies indicated that uptake of infant HIV testing declined after the first two months of life and that misconceptions contributed to poor follow-up [26,27]. The participants of this study confirmed these patterns and emphasised that fear of an HIV positive test result of the infant and fear of unintended disclosure of the HIV status of the parents were major reasons why caregivers delayed or avoided infant testing.

> *“It’s no different with the reality here…parents are afraid that neighbours will know they have HIV. For this reason, mothers do not take their children for testing.”* (Man living with HIV, SSI, Murie)

Participants also linked infant testing gaps to maternal disengagement from care. Babies of women who discontinued ART shortly after delivery were described as being at particularly high risk of missing timely testing and, when needed, early treatment. Several participants also noted that children living with HIV do not always show symptoms in the early stages, which can lead caregivers to delay testing until the child becomes visibly ill. In such cases, testing and treatment were often initiated only once the child’s health had already deteriorated. On the other hand, some caregivers interpreted a negative early infant test result as definitive, leading to the belief that no further HIV testing was necessary. Lack of understanding about the continued risk of HIV transmission during breastfeeding also reduced the perceived need for follow-up testing by some caregivers.

Family dynamics continued to shape decision-making in the postpartum period. While supportive partners and family members could facilitate retention in care and timely infant testing, participants also described cases of partner disengagement after childbirth that hindered follow-up and treatment. They further noted that men and mothers-in-law, despite being key household decision-makers, were rarely directly engaged by HIV programmes, limiting their ability to provide informed support.

Together, these findings highlight the postpartum period as a critical transition during which unresolved disclosure challenges, declining maternal engagement in care, and misconceptions about infant HIV risk converge to weaken continuity of care for both mothers and children. Addressing family-level influences and improving communication about ongoing transmission risks are therefore essential for sustaining engagement in infant testing and maternal treatment during breastfeeding.

#### Participant suggestions for improving HIV service engagement

The study participants proposed several strategies to address barriers identified across the maternal and child health continuum. Before pregnancy, suggestions focused on the need to support couples to discuss sensitive topics such as HIV status and pregnancy intentions. Participants also emphasised the need for stronger engagement of men in family planning and maternal health awareness activities. Some participants suggested that health providers more systematically ask patients receiving ART about pregnancy intentions during routine consultations to create opportunities for preconception counselling.

During pregnancy, participants highlighted the need for better access to information about HIV and treatment. Male participants in particular expressed a desire to be more actively included in educational activities related to HIV, pregnancy, and maternal health.

For the postpartum period, participants proposed strengthened counselling on the risk of mother-to-child transmission during breastfeeding and on the importance of continued ART adherence and infant testing. They emphasised that information should not only target mothers but also fathers and influential family members, particularly mothers-in-law.

## Discussion

This study provides a contextually grounded understanding of how social norms, relationship dynamics, family power structures, misconceptions and lack of information, shape women’s and children’s engagement with HIV services across the maternal and child health continuum in Jangamo and Massinga districts in southern Mozambique. These dynamics unfold within a context where childbearing is closely tied not only to social status but also to lineage continuity and economic security within households. While many barriers identified in this study have been described previously, the SNE approach clarified how these factors operate in practice and revealed specific transition points where engagement in care becomes particularly vulnerable.

Pregnancy emerged as both a motivation and a socially acceptable context for seeking care. Concern for the unborn child encouraged ART adherence and ANC attendance, yet stigma, fear of HIV status disclosure, and unequal power relationships not only with partners but also within the family limited women’s ability to seek care. After delivery, women often lost the social justification that pregnancy had provided for repeated clinic attendance. When HIV status had not been disclosed, ongoing visits to health facilities became harder to explain to partners or family members, contributing to postpartum disengagement, an issue also encountered in PMTCT programmes in other sub-Saharan African countries [28,29,30]. Misinterpretation of early negative infant HIV test results combined with the fact that children living with HIV may not show symptoms in the early stages of infection, reinforced the perception among some caregivers that continued follow-up was unnecessary. In such cases, caregivers sometimes delayed further testing until a child became visibly ill. Similar dynamics have been reported in studies of early infant diagnosis in other sub-Saharan African countries, where mothers’ interpretation of infant test results, in combination with fear of testing outcomes, influenced continued engagement in follow-up care [27,31,32].

The roles of stigma, disclosure challenges, and limited male partner involvement in shaping engagement in PMTCT and HIV care have been widely documented in prior research in Mozambique and other sub-Saharan countries [2,33,22,34]. This study extends existing evidence by clarifying how these factors interact within family structures before and during pregnancy and in the postpartum period to influence specific points of disengagement. The prominent role of mothers-in-law underscores how HIV-related decision-making is embedded in extended household hierarchies, an area that remains underexplored in much of the PMTCT literature [26,35]. Together, these insights move beyond documenting barriers to illuminate the social mechanisms and moments when women and children are most at risk of disengaging from services.

The findings have important implications for HIV programming. Interventions that focus solely on individual knowledge and clinic-based service delivery are unlikely to address the relational and social dynamics that shape care engagement [26,23]. Programmes should place greater emphasis on supporting safe disclosure, strengthening couple communication, and engaging influential family members, particularly male partners and mothers-in-law. Findings related to adolescent pregnancy and conflicting views on contraception also point to the need for age-appropriate strategies that support open dialogue around sexual and reproductive health and rights, helping young people navigate pregnancy prevention and HIV risk before pregnancy occurs. Such efforts are particularly important in settings where gendered power dynamics limit women’s ability to negotiate contraceptive use and other health decisions. Furthermore, the transition from pregnancy to the postpartum period represents a critical vulnerability point, suggesting the need for enhanced counselling and targeted follow-up strategies that retain women and children in care after delivery, including community-based support and differentiated service delivery models that reduce the need for frequent, highly visible clinic visits [36,37]. Clear communication about the meaning of early infant HIV test results and ongoing transmission risk during breastfeeding is also essential to prevent premature disengagement from infant testing and maternal treatment.

This study has several strengths. By using the SNE approach, we were able to systematically build on existing evidence while generating contextually grounded insights into how known barriers operate in practice. Integrating synthesised literature into qualitative data collection facilitated efficient exploration of relevant themes and surfaced locally specific mechanisms that might not have emerged through either a stand-alone literature review or an open-ended qualitative inquiry. The study also engaged diverse stakeholders, enhancing the credibility of the findings. However, findings are based on specific implementation areas of two districts and may not be directly generalisable to other settings. Because narratives were derived from prior literature, discussions may have been shaped by the issues presented, potentially limiting exploration of topics not included in the prompts. In addition, social desirability and the sensitivity of HIV-related topics may have influenced responses.

In conclusion, integrating existing evidence with community perspectives deepened our understanding of the social and relational mechanisms shaping engagement in HIV care for women and children in the study area. By identifying key moments when disengagement is most likely, and the household and community dynamics influencing those transitions, this study highlights opportunities to strengthen HIV programming beyond traditional clinical service delivery. More broadly, the application of the SNE approach illustrates the value of methods that bridge research and implementation by grounding program design in both prior evidence and lived experience.

## Data Availability

All relevant supporting materials for this study have been provided as supplementary files, including the ethical clearance and administrative approval, the study tools, the literature review, the detailed description of the Synthesised Narrative Exploration (SNE) approach, and the analysis table comparing findings from prior literature with newly identified insights. The qualitative data generated and analysed during this study consist of interview and focus group discussion transcripts that contain sensitive information related to HIV status and personal experiences. To protect participant confidentiality, full transcripts are not publicly available. De-identified excerpts supporting the findings are included in the manuscript. Anonymised transcripts may be made available upon request to the corresponding author, subject to approval by the relevant ethics committee. Please note that additional time may be required to fully anonymise the data prior to sharing.

## Acknowledgments

We thank the women and men in Massinga and Jangamo, including adolescents, young people, and people living with HIV, for sharing their perspectives and experiences during the focus group discussions and interviews. We also thank healthcare providers, civil society stakeholders, community leaders and community actors for their insights, as well as participants of the validation meetings whose contributions strengthened the interpretation of the findings.

We also acknowledge the provincial authorities in Inhambane province for endorsing the study, the Ministry of Health for administrative approval, and the Interinstitutional Bioethics Committee of the Faculty of Medicine and Maputo Central Hospital, of Eduardo Mondlane University for providing ethical clearance.

We also thank N’weti staff for supporting participant mobilisation, logistical coordination, and review of the study protocol, tools, and reports.

---

**S1 Appendix. Description of the Synthesised Narrative Exploration Approach**

**S2 Appendix. Description of the literature review**

**S3 Appendix. Study tool**

**S4 Appendix. IRB and MoH approval letters**

**S5 Appendix. Summary of synthesised evidence from prior studies and participant validation and contextualisation**

## References

1. UNAIDS. AIDSinfo — Data on HIV/AIDS. Mozambique 2021 HIV and ART estimates. Joint United Nations Programme on HIV/AIDS (UNAIDS); 2021. Available from: https://aidsinfo.unaids.org/

2. Cuinhane CE, Roelens K, Vanroelen C, Quive S, Coene G. Perceptions and decision-making with regard to pregnancy among HIV-positive women in rural Maputo Province, Mozambique—a qualitative study. BMC Womens Health. 2018;18(1):166. doi:10.1186/s12905-018-0644-7.

3. Gunn JK, Asaolu IO, Center KE, Gibson SJ, Wightman P, Ezeanolue EE, et al. Antenatal care and uptake of HIV testing among pregnant women in sub-Saharan Africa: a cross-sectional study. J Int AIDS Soc. 2016 Jan 18;19(1):20605. doi: 10.7448/IAS.19.1.20605. PMID: 26787516; PMCID: PMC4718968.

4. Callahan T, Modi S, Swanson J, Ng’ambi W, Katirayi L, Chouraya C, et al. Pregnant adolescents living with HIV: what we know, what we need to know, where we need to go. J Int AIDS Soc. 2017;20:21858. doi:10.7448/IAS.20.1.21858.

5. Munguambe K, Boene H, Vidler M, Bique C, Sawchuck D, Potter JA, et al. Barriers and facilitators to healthcare-seeking behaviours in pregnancy in rural communities of southern Mozambique. Reprod Health. 2016;13(Suppl 1):31. doi:10.1186/s12978-016-0141-0.

6. Chagomerana MB, Miller WC, Tang JH, Hoffman IF, Mthiko BC, Phulusa J, et al. Optimizing prevention of HIV mother to child transmission: Duration of antiretroviral therapy and viral suppression at delivery among pregnant Malawian women. PLoS One. 2018;13(4). doi:10.1371/journal.pone.0195033.

7. Cowan JF, Arpadi SM, Ketter N, Rabkin M, Luque AE, Abrams EJ, et al. Early ART initiation among HIV-positive pregnant women in central Mozambique: a stepped wedge randomized controlled trial of an optimized Option B+ approach. Implement Sci. 2015;10:61. doi:10.1186/s13012-015-0249-6.

8. Teasdale CA, Choy M, Tsiouris F, De Gusmao EP, Banqueiro ECP, Couto A, et al. HIV retesting for pregnant and breastfeeding women across maternal child health services in Nampula, Mozambique. PLoS One. 2023 Mar 24;18(3):e0283558. doi: 10.1371/journal.pone.0283558. Erratum in: PLoS One. 2024 Dec 3;19(12):e0315047. doi: 10.1371/journal.pone.0315047. PMID: 36961842; PMCID: PMC10038279.

9. Sherr K, Ásbjörnsdóttir K, A, Crocker J, Coutinho J, de Fatima Cuembelo M, Tavede E, et al. Scaling-up the Systems Analysis and Improvement Approach for prevention of mother-to-child HIV transmission in Mozambique (SAIA-SCALE): a stepped wedge cluster randomized trial. Implement Sci. 2019;14:41. doi:10.1186/s13012-019-0889-z.

10. Lain MG, Chicumbe S, de Araujo AR, Karajeanes E, Couto A, Giaquinto C, et al. Correlates of loss to follow-up and missed diagnosis among HIV-exposed infants throughout the breastfeeding period in southern Mozambique. PLoS One. 2020;15(8). doi:10.1371/journal.pone.0237993.

11. Abou-Setta AM, Jeyaraman MM, Attia A, Al-Inany HG, Ferri M, Ansari MT, et al. Methods for Developing Evidence Reviews in Short Periods of Time: A Scoping Review. PLoS One. 2016;11(12). doi:10.1371/journal.pone.0165903.

12. Lund H, Sørensen J, Bredahl T. Using an evidence-based research approach before a new study is conducted to ensure value. J Clin Epidemiol. 2020;129:92–94. doi:10.1016/j.jclinepi.2020.07.019.

13. Manderson L, Aaby P. An epidemic in the field? Rapid assessment procedures and health research. Soc Sci Med. 1992;35(7):839–850. doi:10.1016/0277-9536(92)90047-X.

14. Macinko J, Almeida C, Klingelhoefer de Sá P. A rapid assessment methodology for the evaluation of primary care organization and performance in Brazil. Health Policy Plan. 2007;22(3):167–177. doi:10.1093/heapol/czm013.

15. Sharma SV, Haidar A, Noyola J, Tien J, Rushing M, Naylor BM, et al. Using a rapid assessment methodology to identify and address immediate needs among low-income households with children during COVID-19. PLoS One. 2020;15(10). doi:10.1371/journal.pone.0240009.

16. McLeroy KR, Bibeau D, Steckler A, Glanz K. An ecological perspective on health promotion programs. Health Educ Q. 1988 Winter;15(4):351–77. doi: 10.1177/109019818801500401. PMID: 3068205.

17. Gombachika B, Fjeld H, Chirwa E, Sundby J, Malata A, Maluwa A (2012). A Social Ecological Approach to Exploring Barriers to Accessing Sexual and Reproductive Health Services among Couples Living with HIV in Southern Malawi. International Scholarly Research NetworkISRN Public HealthVolume 2012, Article ID 825459, 13 pagesdoi:10.5402/2012/825459

18. Dyson YD, Mobley YP, Harris G, Randolph SD. Using the social-ecological model of HIV prevention to explore HIV testing behaviors of young Black college women. Journal of the Association of Nurses in AIDS Care. 2018;29(1):53–59. Available from: https://pubmed.ncbi.nlm.nih.gov/29274654/

19. World Health Organization. Packages of interventions for maternal, newborn and child health: continuum of care. WHO/FCH/10.06. Geneva: World Health Organization; 2010.

20. Kerber KJ, de Graft-Johnson JE, Bhutta ZA, Okong P, Starrs A, Lawn JE. Continuum of care for maternal, newborn, and child health: from slogan to service delivery. The Lancet. 2007;370(9595):1358–1369

21. Hilliard S, Gutin SA, Dawson Rose C. Messages on pregnancy and family planning that providers give women living with HIV in the context of a Positive Health, Dignity, and Prevention intervention in Mozambique. Int J Womens Health. 2014 Dec 12;6:1057–67. doi: 10.2147/IJWH.S67038.

22. Audet CM, Blevins M, Chire YM, Vaz LM, Antonio E, Gouveia ML, et al. Engagement of Men in Antenatal Care Services: Increased HIV Testing and Treatment Uptake in a Community Participatory Action Program in Mozambique. AIDS Behav. 2016;20(9):2090–2100. doi:10.1007/s10461-016-1341-x.

23. Workneh NG, Kevany S. Maternal Health Service Disparities Across Incomes and Implications on Prevention of Mother-to-Child Transmission Service Coverage: Current Context in Sub-Saharan Africa. J Public Health Afr. 2016;7(2):402. doi:10.4081/jphia.2016.402.

24. Llenas-Garcia J, Wikman-Jorgensen P, Hobbins M, Mussa MA, Ehmer J, Keiser O, et al. Retention in care of HIV-infected pregnant and lactating women starting ART under Option B+ in rural Mozambique. Trop Med Int Health. 2016;21(8):1003–1012. doi:10.1111/tmi.12728.

25. Stover KE, Shrestha R, Tsambe I, Mathe PP. Community-Based Improvements to Increase Identification of Pregnant Women and Promote Linkages to Antenatal and HIV Care in Mozambique. J Int Assoc Providers AIDS Care. 2019 Jan-Dec;18:2325958219855623. doi: 10.1177/2325958219855623.

26. De Schacht C, Lucas C, Mboa C, Gill M, Macasse E, Dimande S, et al. Access to HIV prevention and care for HIV-exposed and HIV-infected children: a qualitative study in rural and urban Mozambique. BMC Public Health 2014, 14;1240. doi:10.1186/1471-2458-14-1240

27. Sibanda EL, Weller IV, Hakim JG, Cowan FM. The magnitude of loss to follow-up of HIV-exposed infants along the prevention of mother-to-child HIV transmission continuum of care: a systematic review and meta-analysis. AIDS. 2013;27(17):2787–2797.

28. Knettel BA, Cichowitz C, Ngocho JS, Knippler ET, Chumba LN, Mmbaga BT, et al. Retention in HIV Care During Pregnancy and the Postpartum Period in the Option B+ Era: Systematic Review and Meta-Analysis of Studies in Africa. J Acquir Immune Defic Syndr. 2018 Apr 15;77(5):427–438. doi: 10.1097/QAI.0000000000001616.

29. Hodgson I, Plummer ML, Konopka SN, Colvin CJ, Jonas E, Albertini J, et al. A systematic review of individual and contextual factors affecting ART initiation, adherence, and retention for HIV-infected pregnant and postpartum women. PLOS ONE. 2014;9(11):e111421. 10.1371/journal.pone.0111421

30. Geldsetzer P, Yapa HM, Vaikath M, Ogbuoji O, Fox MP, Essajee SM, et al. A systematic review of interventions to improve postpartum retention of women in PMTCT and ART care. Journal of the International AIDS Society. 2016;19(1):20879. 10.7448/IAS.19.1.20879

31. Adeniyi VO, Thomson E, Goon DT, Ajayi IA. Disclosure, stigma of HIV positive child and access to early infant diagnosis in the rural communities of OR Tambo District, South Africa: a qualitative exploration of maternal perspective. BMC Pediatr 15, 98 (2015). 10.1186/s12887-015-0414-8

32. Etoori D, Renju J, Reniers G, Ndhlovu V, Ndubane S, Makhubela P, et al. ‘If the results are negative, they motivate us’. Experiences of early infant diagnosis of HIV and engagement in Option B+. Global Public Health. 2021: 16(2), 186–200. Doi:10.1080/17441692.2020.1795220

33. Mbonu NC, van den Borne B, De Vries NK. Stigma of people with HIV/AIDS in Sub-Saharan Africa: a literature review. J Trop Med. 2009;2009:145891. doi:10.1155/2009/145891.

34. Wesevich A, Mtande T, Saidi F, Cromwell E, Tweya H, Hosseinipour MC, et al. Role of male partner involvement in ART retention and adherence in Malawi’s Option B+ program. AIDS Care. 2017;29(11):1417–1425. doi:10.1080/09540121.2017.1308464.

35. Banke-Thomas OE, Banke-Thomas AO, Ameh CA. Factors influencing utilisation of maternal health services by adolescent mothers in low- and middle-income countries: a systematic review. BMC Pregnancy Childbirth. 2017;17:65. doi:10.1186/s12884-017-1246-3.

36. Pfeiffer JT, Napúa M, Wagenaar BH, Chale F, Hoek R, Micek M, et al. Stepped-Wedge Cluster Randomized Controlled Trial to Promote Option B+ Retention in Central Mozambique. J Acquir Immune Defic Syndr. 2017;76(3):217–225. doi:10.1097/QAI.0000000000001490.

37. Kawakyu N, Nduati R, Munguambe K, Coutinho J, Mburu N, DeCastro G, et al. Development and Implementation of a Mobile Phone–Based Prevention of Mother-To-Child Transmission of HIV Cascade Analysis Tool: Usability and Feasibility Testing in Kenya and Mozambique. JMIR Mhealth Uhealth. 2019;7(5). doi:10.2196/13963.

